# The influence of ensemble size and composition on the performance of combined real-time COVID-19 forecasts

**DOI:** 10.1101/2025.08.09.25331484

**Authors:** Friederike Becker, Katharine Sherratt, Nikos Bosse, Sebastian Funk

## Abstract

During infectious disease outbreaks, short-term forecasts can play an important role for both decision makers and the general public. While previous research has shown that combining individual forecasts into an ensemble improves accuracy and consistency, practical guidance for organisers of multi-model prediction platforms on how to construct an ensemble has been scarce. In particular, it is not entirely clear how ensemble performance relates to the size of the underlying model base, a relevant question when relying on voluntary contributions from modelling teams that face competing priorities. Furthermore, the exact composition of an ensemble forecast may influence its performance. Ensembles can either include all models equally or, alternatively, discriminate based on past performance or other characteristics.

Using data from the European COVID-19 Forecast Hub we investigated these questions, with the aim of offering practical guidance to organisers of multi-model prediction platforms during infectious disease outbreaks. We found that including more models both improved and stabilized aggregate ensemble performance, while selecting for better component models did not yield any particular advantage. Diversity among models, whether measured numerically or qualitatively, did not have a clear impact on ensemble performance.

These results suggest that for those soliciting contributions to collaborative ensembles there are more obvious gains to be made from increasing participation to moderate levels than from optimising component models.

## 1 Introduction

Epidemic forecasting can be a useful tool during infectious disease outbreaks, reducing short-term uncertainty and supporting planning and resource allocation (Grieve et al. 2023). Repeated studies have demonstrated increased forecast accuracy from the combination of multiple model predictions into an ensemble. This result has been noted since the 1960s (Bates et al. 1969), and repeatedly observed in outbreak forecasting across multiple pathogens and outbreaks over time (e.g., Viboud et al. 2018; McGowan et al. 2019; Cramer et al. 2022; Sherratt et al. 2023). It also motivates calls for continued investment in large-scale modelling collaborations producing ensemble forecasts (Reich et al. 2022).

Meanwhile, collaborative ensembles are typically “ensembles of opportunity” (Knutti et al. 2010), combining across a non-systematically selected, and potentially fluctuating, sample of contributing models with limited treatment of model interdependence or performance in ensemble construction. Therefore, even while ensemble predictions tend to outperform the majority of individual models, it is important to assess whether this performance is robust to different compositions of contributing models. Evaluating contributing factors for creating ensemble outperformance can also support the design of future collaborative modelling projects.

Contributing factors to ensemble performance may be the number of member component models, the predictive performance of those individual models, and their diversity. Previous work analysing US influenza and COVID-19 forecasts has characterised the impact of the number of component models (Fox et al. 2024). This found that among a random selection of models, including more models improved ensemble performance, particularly in the range of up to four models, while also decreasing the variability of ensemble performance.

Studies in various fields have discussed and empirically shown the value of including a diverse set of forecasts in an ensemble (e.g., Lamberson et al. 2012; Batchelor et al. 1995). Past work on forecasting infectious diseases has investigated differences in forecast performance from individual methodological types (Viboud et al. 2018; Johansson et al. 2019), but has not identified the impact of combining across these in resulting ensemble performance.

Apart from diversity, an ensemble might identify or weight its potential members based on their individual performance. In many applications, equally weighted ensembles are not outperformed by approaches that assign model weights based on individual past performance (Claeskens et al. 2016). In related analyses to ours, Bracher et al. (2021b) found no systemic benefits of a weighted ensemble using data from Germany and Poland, while Ray et al. (2023) have conversely found a weighted approach to be beneficial in forecasting COVID-19 Deaths, based on data from the US.

In this work we explore the impact of these factors in the context of the European COVID-19 Forecast Hub, a collaborative multi-model project set up in 2021 to inform the response to the COVID-19 pandemic across Europe. We particularly focus on the influence of ensemble size (number of contributing models) and diversity, and implications for ensemble constructions including attempts to select best performers.

## 2 Methods

### 2.1 Scope

We used forecast data from the European COVID-19 Forecast Hub (Sherratt et al. 2022), a coordinated and collaborative forecasting effort instigated by the European Centre for Disease Prevention and Control (ECDC) in 2021. The Hub collated weekly forecasts from independent modelling teams for COVID-19 Deaths and Cases, for one to four weeks into the future and across 32 European countries. Forecasts were submitted in a probabilistic and standardized format, via 23 predictive quantiles, that is, 11 central prediction intervals plus a median forecast.

All forecasts were evaluated against ground truth data collated for public use by Johns Hopkins University (JHU), with weekly incident Cases and Deaths derived from the daily cumulative counts reported by JHU by taking differences, as was done in the data set used as ground truth for the Hub.

During the period studied here, the Hub included a baseline model (last value carried forward, with uncertainty stemming from previous weekly differences of the series, analogous to Cramer et al. 2022) and an official Hub ensemble, which aggregated all eligible component forecasts initially via the mean and, later, the median function, and was used by the ECDC in communication about the development of the pandemic.

In our analysis, we generally compared the performance of alternatively constructed ensembles to that of a “Hub-replica” ensemble, which aggregated all available models via the median function at all forecast dates. This removed the structural break that the switch of aggregation method presented in the official Hub ensemble data.

We constructed alternative ensembles to investigate the impact of changing the composition of the ensemble forecast, and thereby relied critically on the size of the available model base. We thus limited ourselves to a subset of the Hub data that generally saw higher numbers of individual submissions from the independent modelling teams, while still retaining some generality. Concretely, we used forecasts that were issued for Germany, Poland, the Czech Republic, the United Kingdom and France, for both Deaths and Cases, between Mid-March 2021 and Mid-March 2022. This gave us 52 forecast dates in total. Figure 1 displays the number of models that were available for these locations and both target series. Submission levels were higher during the summer and fall months of 2021, and the highest levels of participation were generally observed in Germany and Poland.

**Figure 1.**
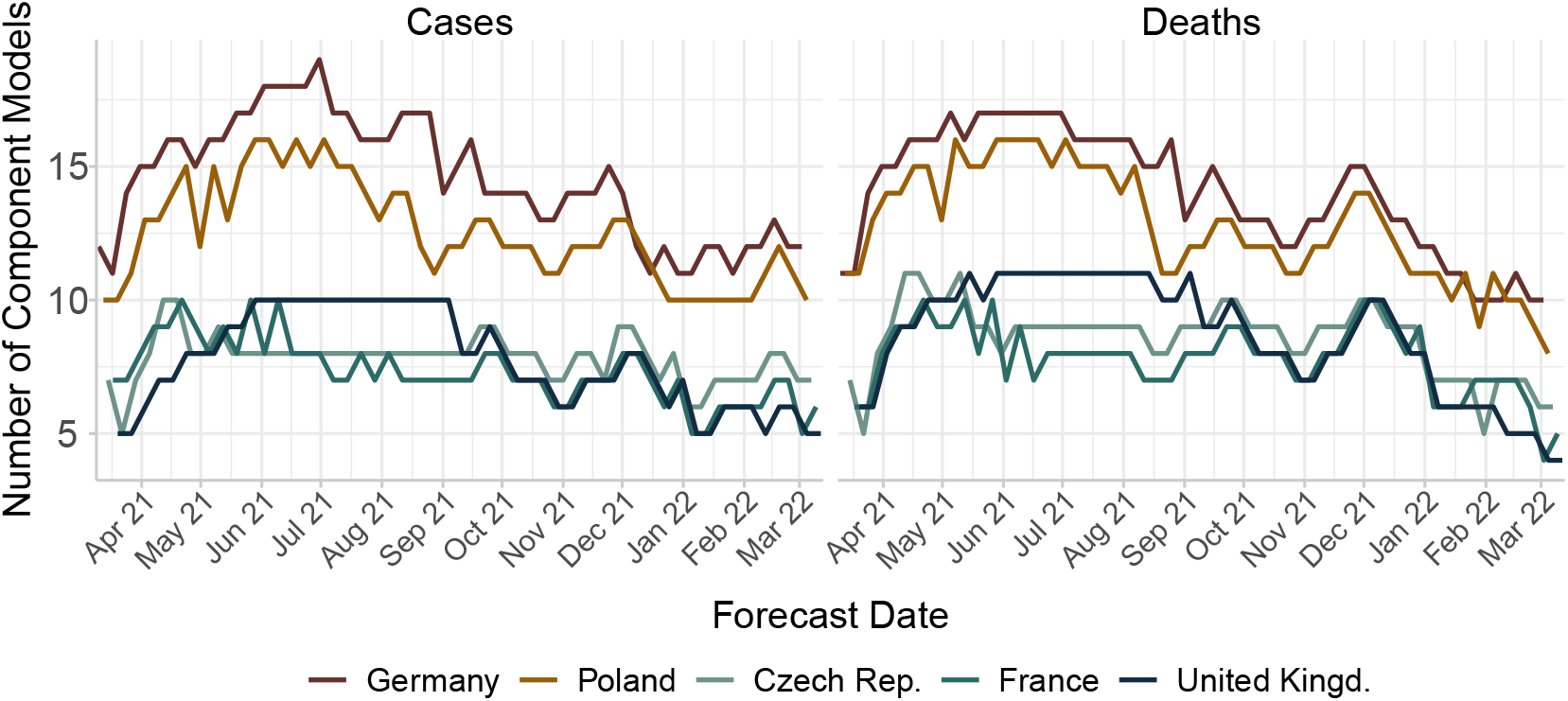
Number of available component models for each location and the two target series, COVID-19 Cases and Deaths.

For our study, we only considered “full” forecasts, that is, those that were made for all horizons up to 4 weeks ahead and all 23 quantile levels. We generally focused on communicating results for the 1- and 2-week horizon. This choice is due to conciseness, and where appropriate, we refer to results for all horizons in the appendix.

### 2.2 Predictive performance

Similarly to previous analyses on the US and European COVID-19 Forecast Hubs (e.g., Sherratt et al. 2023; Cramer et al. 2022), we scored all forecasts with the weighted interval score (WIS, Bracher et al. 2021a), a proper scoring rule designed for forecasts that are submitted in an interval format. The WIS is negatively oriented - that is, smaller values correspond to better forecasts.

In the first part of our analysis, we constructed alternative ensembles that each made forecasts on distinct subsets of the training sample, see the description of the recombination process in section 2.3. As a result, individual ensembles’ average WIS values were not directly comparable. We addressed this issue of intermittent submissions with so-called pairwise comparisons (Cramer et al. 2022). The Hub-replica ensemble was used as the benchmark model to scale average pairwise scores against. We thereby obtained a concise measure of performance, scaled relative skill, which can be interpreted as follows: a value *>* 1 means that the respective alternative ensemble performed worse than the Hub-replica ensemble, while a value *<* 1 means that it performed better.

In the second part of our analysis, we constructed ensembles that selected for better performers on a rolling basis. The resulting ensembles were available for all forecast dates and we thus obtained a comparable measure of performance by dividing the WIS of the given alternative ensemble with that attained by the Hub-replica ensemble. Again, a value *>* 1 means that the respective alternative ensemble performed worse than the Hub-replica ensemble, and vice versa.

### 2.3 Construction of ensembles of different size

For the first part of our analysis, we constructed alternative ensembles by recombining from the total set of models, which is distinct for each combination of target series *j* (Cases and Deaths) and forecast location (country) *i*. For each location-target, and for each *k*, we thereby obtained 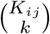proposed ensembles, where *K*_*ij*_ is the total number of models that have at some point submitted a forecast for target *j* and location *i*.

The available forecast data was characterized by intermittent submission (not all models were submitted for all dates, see variability in available models in Figure 1), which posed some issues for scoring forecasts, see subsection 2.2. We thus imposed some availability thresholds on individual models and the recombined ensembles. Individual models were considered as eligible for recombination if forecasts were available for at least half of the study period. We thus obtained 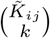 suggested recombined ensembles, each of which was then considered eligible to be scored as an alternative ensemble if all models were *jointly* available for at least half of the study period. Final candidate ensembles were then aggregated separately with the median and mean functions.

Recombined ensembles were additionally classified as being either “homogeneous” (all included component models were of the same general modelling type, either “mechanistic”, “semi-mechanistic” or “statistical”) or “heterogenous” (included component models were of two or more different types). Component models were classified based on metadata provided by the teams, following the same classification as in Sherratt et al. (2025), which provides further details on the process. Using this classification of ensembles we could investigate whether those ensembles that incorporated diverse modelling approaches tended to perform better than those that relied on a single model type.

We took random samples (at least 75 percent of recombined ensembles) if a location-target combination had such a large number of component models that the computational burden of running pairwise comparisons for the whole recombination set was too high. We initiated via several different random seeds and thereby checked that results were not meaningfully affected by this step.

### 2.4 Construction of best-performer ensembles

For the second part of our analysis, we investigated if and how it may be beneficial to compose an ensemble by selecting for better recent performers, possibly in conjunction with additionally weighting models based on their recent performance.

To this end, we used a rolling window approach at each forecast date to select the *k* best recent performers, based on 1- and 2-week horizon WIS values for the respective location and target series in the past four weeks. To best reflect the original reals-time setting, we only included forecasts that had already realized at the respective forecast date, thereby excluding the most recent 2-week forecast. For the number of models selected into the ensemble, we considered *k* = 3, 5, 8, 10. The *k* chosen models were then aggregated with two methods, once equally and once via their inverse WIS value in the past four weeks (since the WIS is negatively oriented, better recent performers thus received a higher weight).

While the Hub favoured the median due to its robustness to potential outlying forecasts, we additionally investigated mean aggregation here. It is possible that the mean could perform better in the given setting, where we selected for better recent performers and thus potentially excluded those models that might be prone to outlying predictions.

Given that our analysis was entirely retrospective, we refrained from excessively tuning the window size to the point where it would give the most favourable in-sample results. We therefore chose a similar value as Bracher et al. (2021b) in a related analysis, and slightly increased this to four weeks because we had access to a larger dataset.

Ray et al. (2023) and Sherratt et al. (2023) conducted similar analyses on the European Hub data. However, the former did not treat locations separately and thus did not account for the considerable difference in model availability by location, while the latter did not preselect models before weighting them.

### 2.5 Computation

All analyses were conducted using the statistical software R. The code for this analysis can be found here: https://github.com/fredbec/eurohub-prt/. All scoring and performance evaluation was done with the scoringutils R package (Bosse et al. 2024).

## 3 Results

### 3.1 Number of models contained in ensemble

Relative WIS values for recombined ensembles aggregated with the median are shown in Figure 2, respectively for Cases and Deaths and the two forecast horizons. For each set of recombined ensembles of size *k*, we show the median scaled relative skill, as well as the min-max and 5% − 95% quantile range.

**Figure 2.**
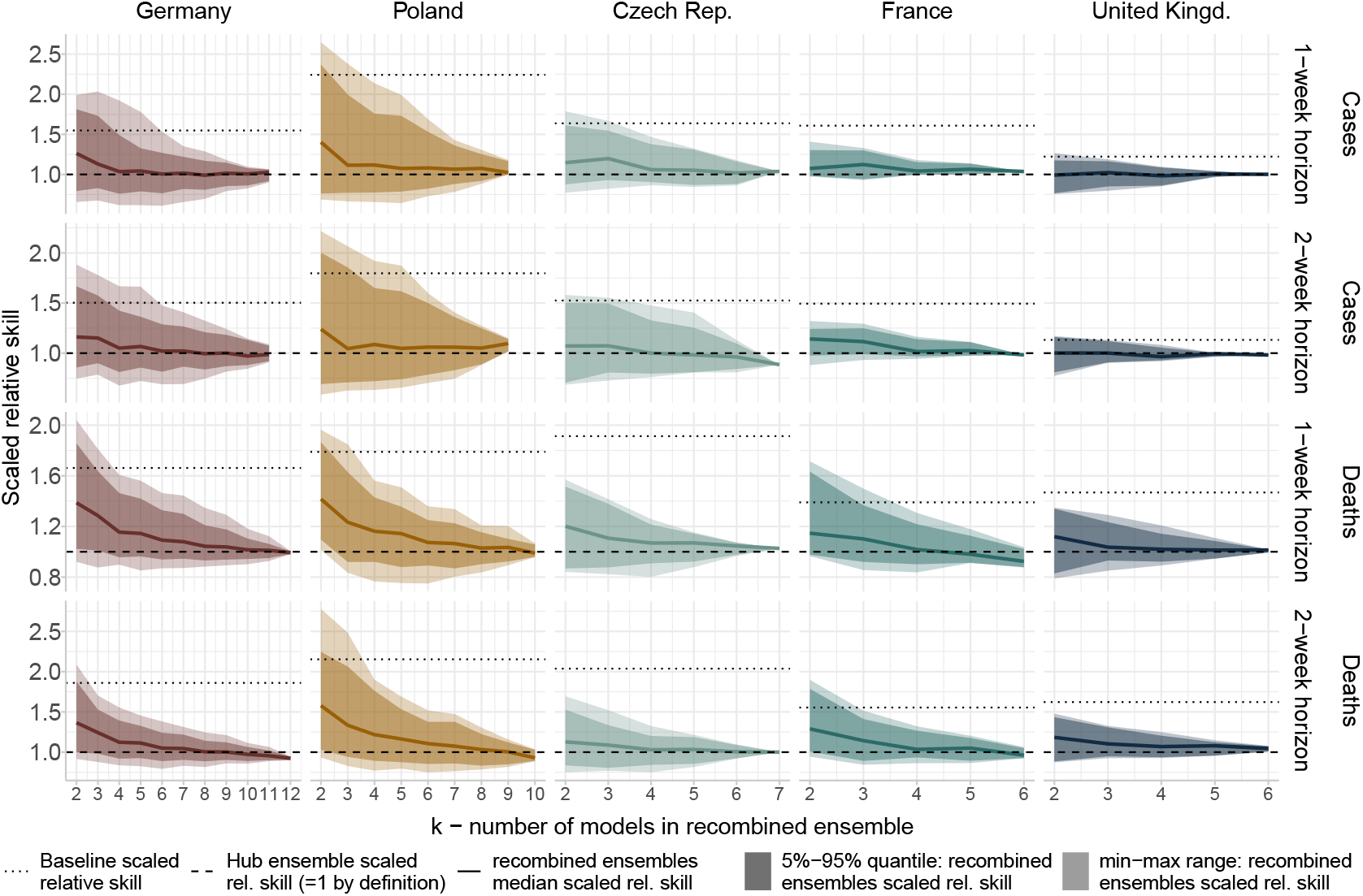
Ensemble recombination results for COVID-19 Cases and Deaths, using data from the European COVID-19 Forecast Hub. Models were recombined from the full set of available forecast models to create alternative ensembles of size *k*, which were scored via the weighted interval score and pairwise comparisons, and subsequently scaled against the performance of the full Hub-replica ensemble. Recombined ensembles were aggregated with the median function. Curves differ by length, depending on the size of the available underlying model base. For Germany, recombined ensembles were sampled with 75% frequency due to computational costs - to check robustness of results, the analysis was separately rerun with different random seeds.

For both target series and all horizon-location combinations, the benefits of ensembling component forecasts were apparent, with values of scaled relative skill generally decreasing with a larger number of component models in the recombineds ensembles. With few exceptions, recombined ensembles with 4 members or more outperformed the baseline model. For most series, the largest decreases in median scaled relative skill tended to occur between 2 and 4 component models, after which performance typically either plateaued or improved more slowly. In particular, larger digit percentage decreases in median scaled relative skill across all recombined ensembles were typically limited to smaller values of *k*, see Supplemental Figure 6, while percentage changes for larger *k* tended to stay closer to zero. In some instances, moderate improvements continued to higher values of *k*, e.g., for Poland Deaths.

Spread in relative performance also considerably decreased with *k*, suggesting that including more models may remove the need to select component models that are particularly beneficial for ensemble performance. Some of this effect is likely attributable to the recombined ensembles simply becoming more similar in their composition. However, the decrease in performance variability was still apparent in the range of smaller *k* values, where there were still recombined ensembles that have zero overlap in the models that they included.

Overall, differences between the two forecast horizons and the two target series appeared small, with all curves largely following the same pattern. Between the different locations, the number of available component models was a major factor in the different forms of the curves.

Additionally, Figure 3 shows the range in scaled relative skill for recombined ensembles whose component members are all of the same model type (“homogeneous”) or of two or more different model types (“heterogeneous”). While sample sizes were small for homogeneous models in particular, the performance ranges for the two groups were similar, and in particular not clearly in favour of heterogeneous ensembles, especially for ensembles with more than two members. When considering a direct numerical measure of distance between component model predictions in each recombined ensemble, see Supplemental Figure 7, there also appeared to be no clear relationship between diversity in an ensemble and that ensemble’s performance.

**Figure 3.**
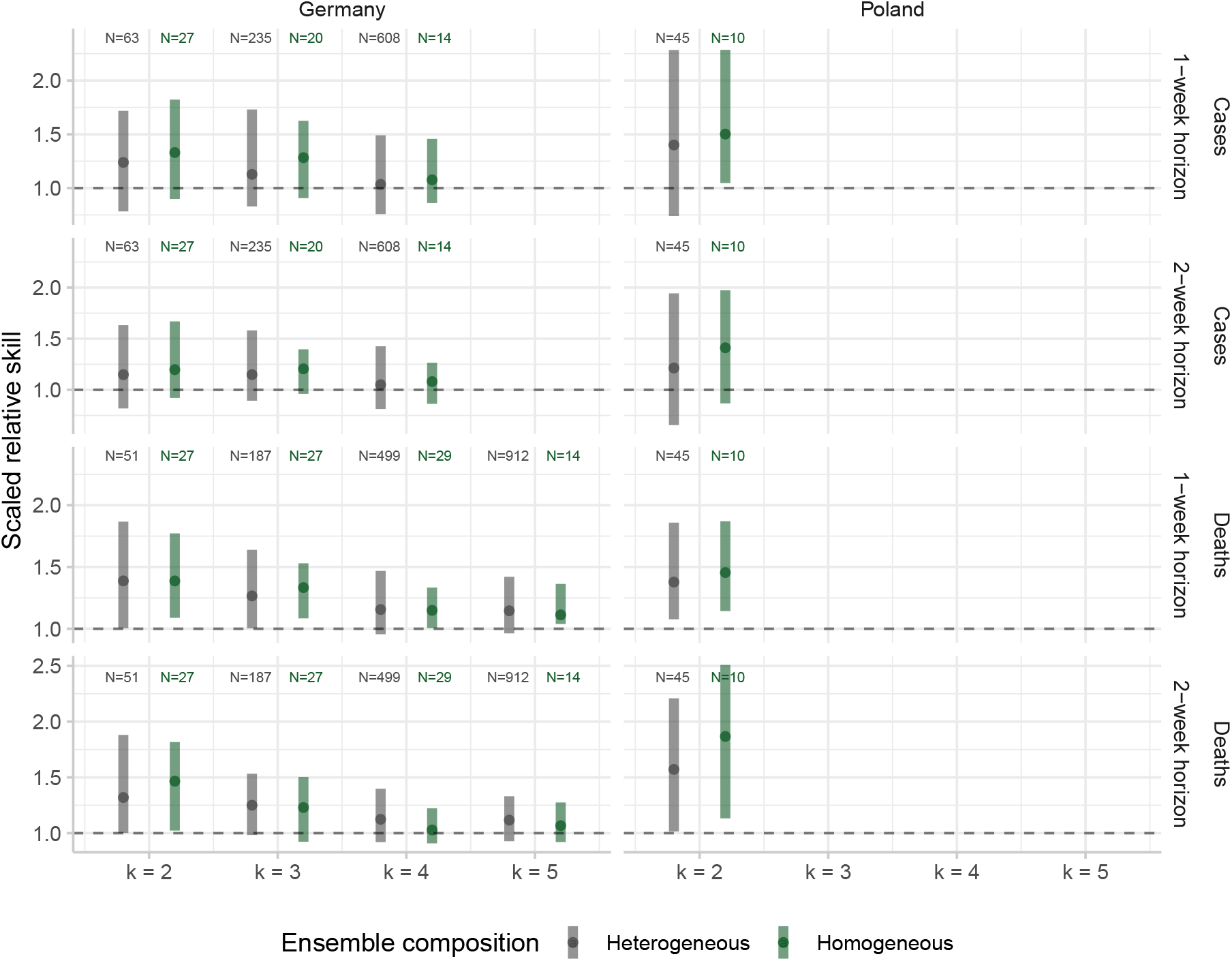
Same data as Figure 2, with ensembles split up by types “homogeneous” (all component models in the recombined ensemble were of the same model type: either mechanistic, semi-mechanistic, or statistical) and “heterogeneous” (component models in the ensemble were of two or more different model types). Shown are 5% to 95% quantile ranges for scaled relative skill (shaded areas) and median scaled relative skill (points). Data is only shown for combinations of location, target series and k for which there were at least 10 recombined ensembles of each type. The number of available recombined ensembles is indicated at the top of each plot, via ‘N’.

Any “jumpiness” or kinks in the relative skill curves in Figure 2 could be attributed to the median aggregation method, which repeatedly switched from selecting the midpoint prediction (uneven number of component models) and averaging between the two midpoint predictions (even number of component models). Particularly in the range of smaller *k*, this did have an effect on results. Supplemental Figure 5 shows the results from the same experiment with mean aggregation. Overall, the results were very similar, with the curves following the same patterns but appearing noticeably smoother.

### 3.2 Best-performers ensemble

Figure 4 shows the average WIS values of the selection ensembles, relative to the Hub-replica benchmark ensemble. Especially when averaged across all locations (leftmost column in both plots), relative WIS values stayed close to one in most cases, indicating no substantial performance gains (or losses) when pre-selecting for recent better performers, whether or not they are subsequently weighted.

**Figure 4.**
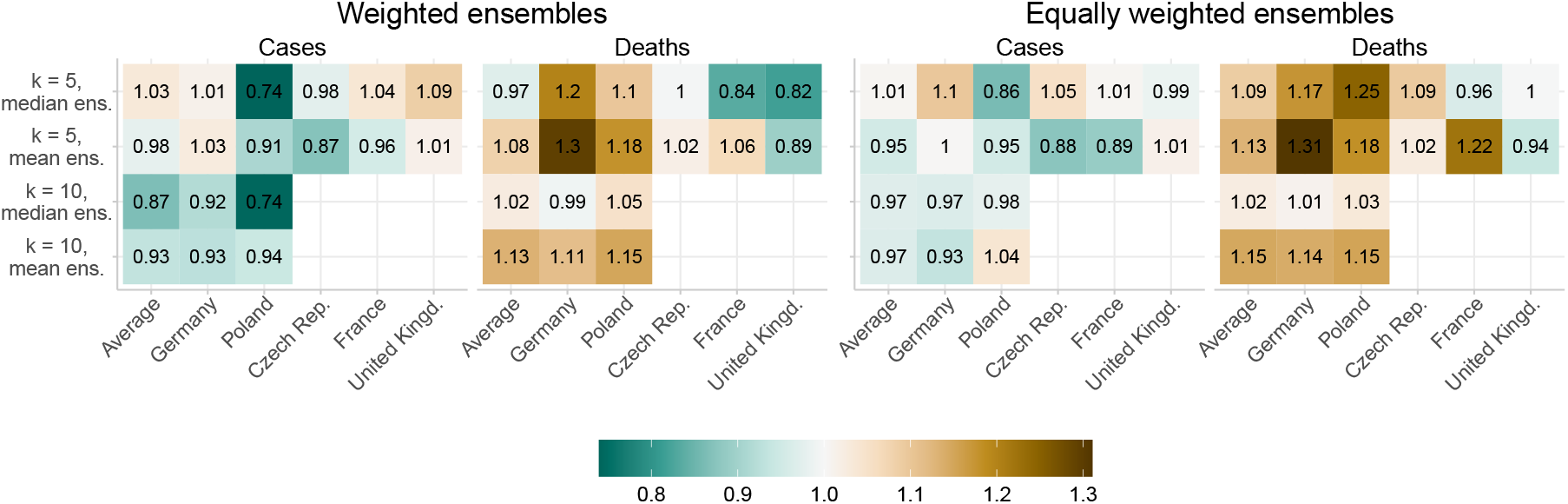
Average WIS of the (weighted) selection methods, divided by the full Hub-replica ensemble, for the 1- and 2-week forecast horizons. The selection method reduced the set of component forecasters to the number of *k* best recent performers - different aggregation methods were then applied to obtain the ensemble (weighted or unweighted mean or median). The “Average” location refers to the setting where scores were averaged across all locations. Relative scores are missing whenever a location’s model base was not large enough to support the analysis.

In fact, for any given aggregation method, the selection ensemble tended to both outperform and be outperformed by the benchmark in some locations, suggesting that sample variation might be a key factor underlying the relative performance scores. For Deaths, relative scores were often substantially above one, indicating that the full unweighted benchmark ensemble was favoured. For Cases, relative scores tended to stay close to one, indicating no clear favourite. Differences were overall small between the weighted and unweighted approaches, with a slight preference for the weighted ensembles. Results for all horizons are shown in Supplemental Figure 8, showing that scores tended to worsen slightly when additionally including forecast horizons of three and four weeks. A consistent improvement in performance seemed to be limited to the Cases series in Poland, see also the boxplots showing spread of relative WIS values over all forecast dates in Supplemental Figure 9.

To summarise, improvements in performance were moderate at best, and were not consistent for any aggregation method across all locations and both target series. Supplemental Figure 9 shows WIS value spreads decreasing when more models were selected into the ensemble, suggesting ensemble size, rather than selecting for better component models, to be the more important factor when aiming to stabilize ensemble performance.

## 4 Discussion

Our analysis of the number of models contained in an ensemble showed that including more models both on average improved and stabilized aggregate ensemble performance. These improvements however tended to taper off or become more gradual after 2 to 4 models. We found no strong connection between an ensemble’s diversity and its performance. This was the case both when assessing diversity numerically and when doing so qualitatively in terms of classifying modelling approaches.

While results showed that, even with a low number of component models, some recombined ensembles performed better than an ensemble that included all available models, it is as yet unclear how these sets of models could be identified in (pseudo-)real time. In fact, there were some location-target combinations where (almost) none of the smaller ensembles managed to outperform the full Hub ensemble, making the latter the more stable choice. The reduction in performance spread further suggested that there may be less need for model selection once an ensemble includes a moderate number of component models.

In particular, the second part of the analysis has shown that selecting for recent better performers did not consistently improve performance and only rarely gave a substantial advantage over a larger equally-weighted median ensemble. This might be due to the non-stationarity of model performance, making past performance of a model an unreliable predictor of current or future performance. On the other hand, more sophisticated aggregation methods might leverage individual contributions more successfully than our attempts to select models on a binary basis and additionally weighting by inverse past scores.

Previous analyses of multi-model infectious disease forecasting efforts have obtained similar results to ours. Fox et al. (2024) identified a stabilizing effect of larger ensembles, while also reporting diminishing returns in performance for more than four models. In a nowcasting context, Amaral et al. (2025) did not find consistent improvements from selection and weighting schemes compared to a full unweighted ensemble, while similarly reporting a stabilization effect of larger ensembles. Ray et al. (2023) employed a more sophisticated weighting scheme than that used in the context of this paper, and found improvements for only one of the two considered data series, COVID-19 Deaths in the United States. In a broader realm, our finding may represent an instance of the “forecast combination puzzle”, that is, the phenomenon that weight estimation often does not improve ensemble performance, see, e.g., Claeskens et al. (2016) for a theoretical discussion.

While most of these existing analyses were limited to a single location, our work extends to several locations and two target series, in a setting characterised by intermittent submission and, at times, a small number of available models.

Extensions of our analysis could include the application of more sophisticated weighting schemes. Likewise, a selection scheme could focus on particularly well performing subgroups of models rather than individual models, as in Fox et al. (2024).

In the absence of consistently better performing component models and/or evidence of more sophisticated aggregation methods, the practical advice for organisers of multi-model prediction platforms thus remains to collate a moderate number of models and aggregate them indiscriminately via the unweighted median function.

## Data Availability

The code for this analysis can be found in the following GitHub repository: https://github.com/fredbec/eurohub-prt
The data used in this analysis stems from the European COVID-19 Forecast Hub. The data is available through https://doi.org/10.5281/zenodo.7669867.

https://github.com/fredbec/eurohub-prt

## 5 Acknowledgements

SF and KS were supported by Wellcome (210758/Z/18/Z).

## A Ensemble Size

**Figure 5.**
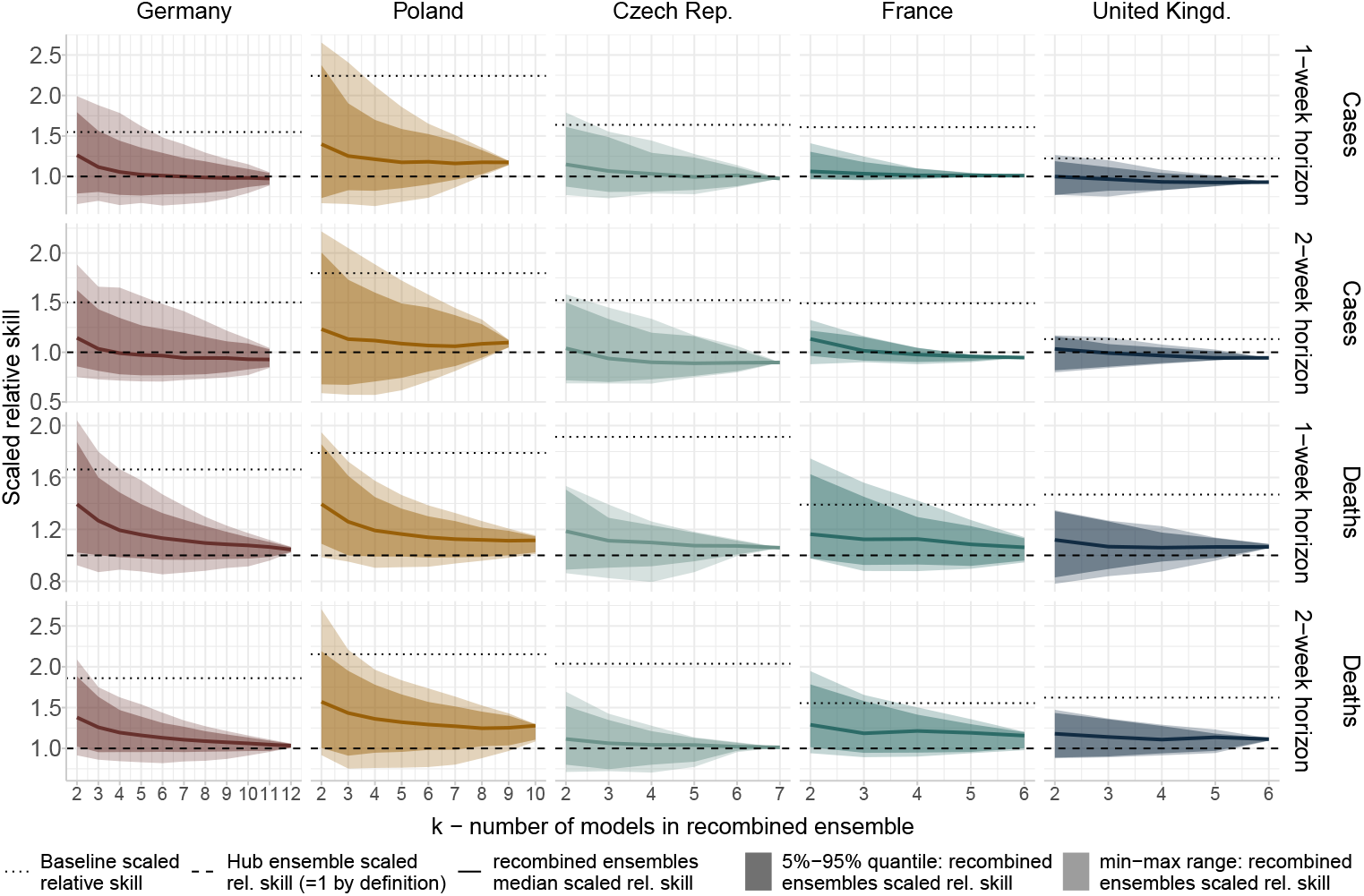
Same as Figure 2, but using the mean function to aggregate recombined ensembles.

**Figure 6.**
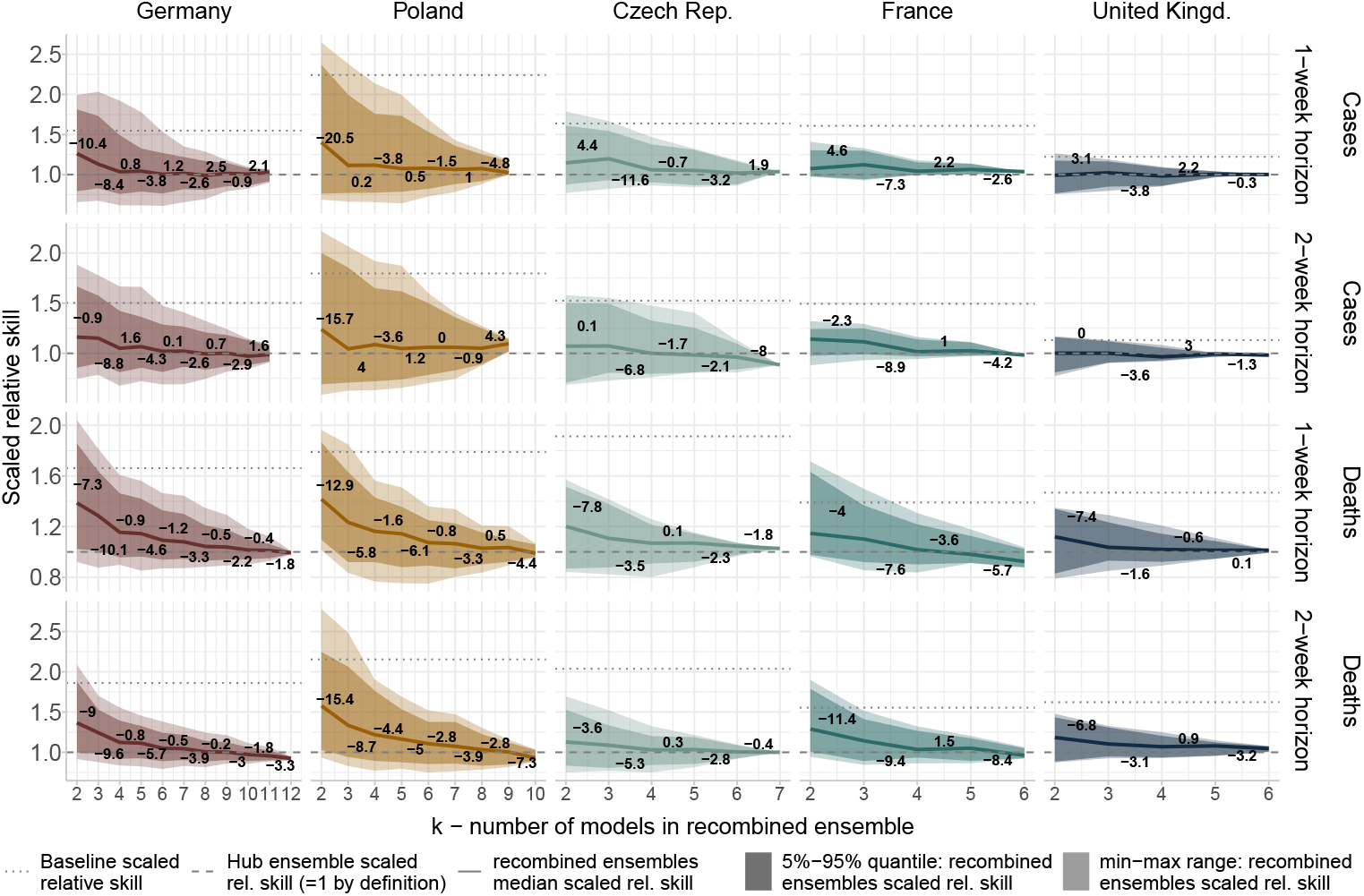
Same as Figure 2, with additionally indicated percentage changes of median scaled relative skill across all recombined ensembles. For instance, a value of, e.g., -9 shown between *k* = 2 and *k* = 3 indicates a decrease of 9% for median scaled relative between *k* = 2 and *k* = 3.

## B Ensemble diversity

**Figure 7.**
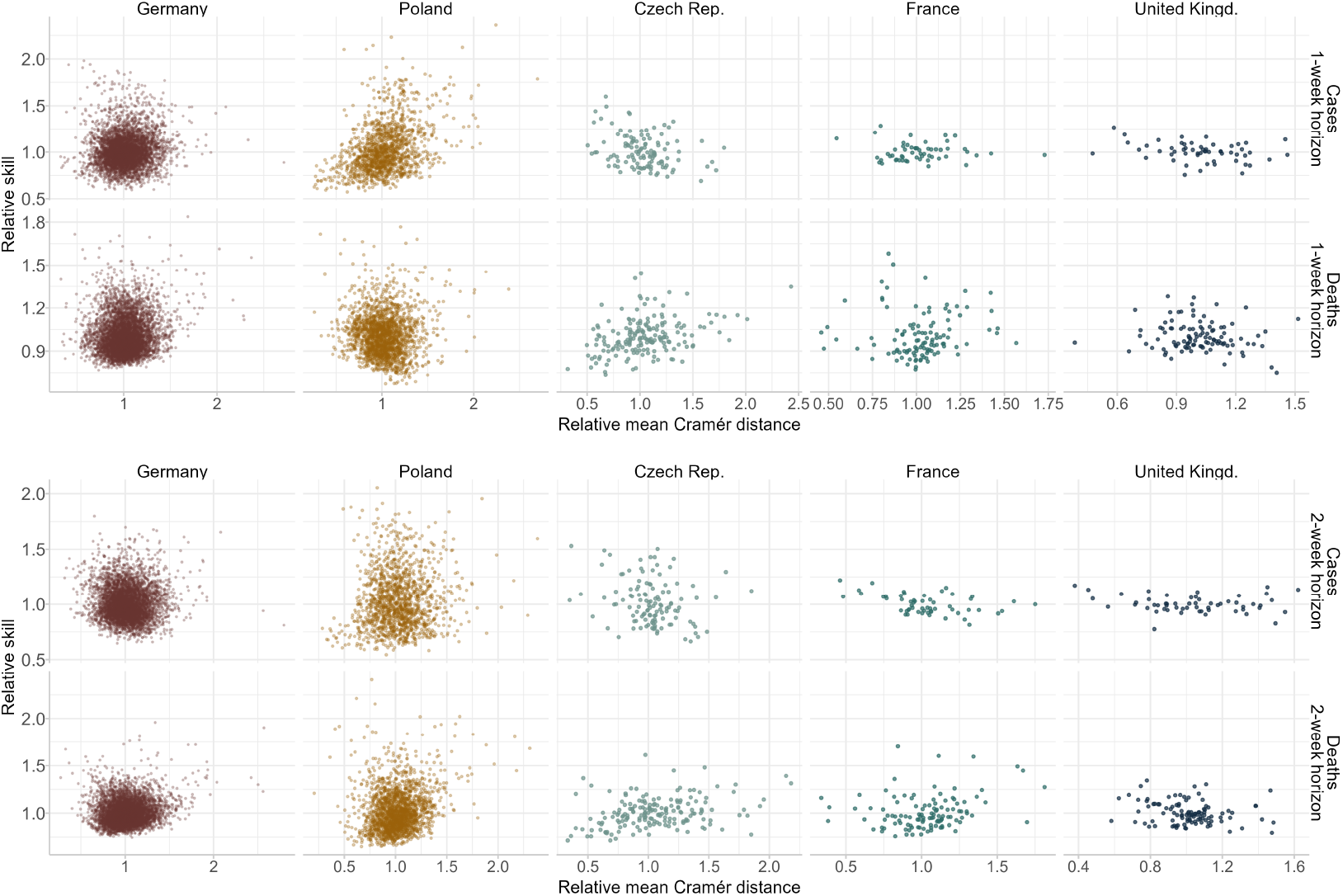
Scaled relative skill, plotted against relative mean pairwise Cramér distance, for all recom-bined ensembles. The Cramér distance is a measure of dissimilarity between probability distributions, see e.g. Resin et al. (2024) for a quantile-based representation. Each point represents one recombined ensemble. Note that points’ size and alpha values (translucency of points) is slightly adjusted for locations with larger numbers of recombined ensembles (Poland and Germany) to improve legibility.

## C Best-performers ensemble

**Figure 8.**
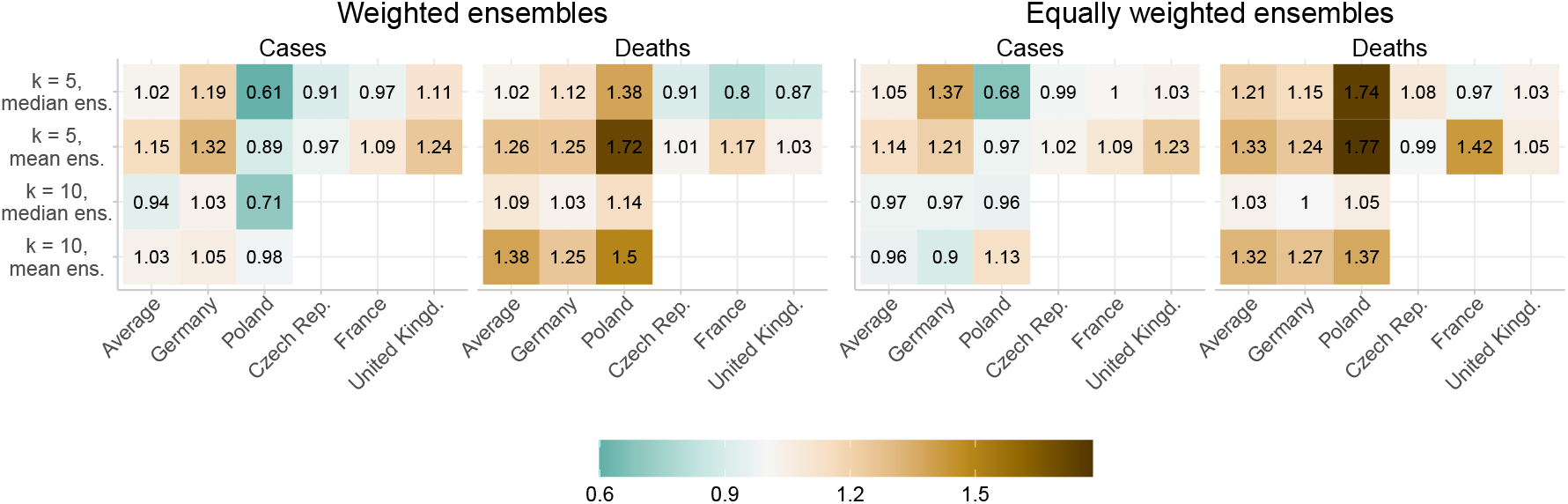
Same as Figure 4, but for all forecast horizons. Note the change in colour scale to accom-modate a larger range of relative scores.

**Figure 9.**
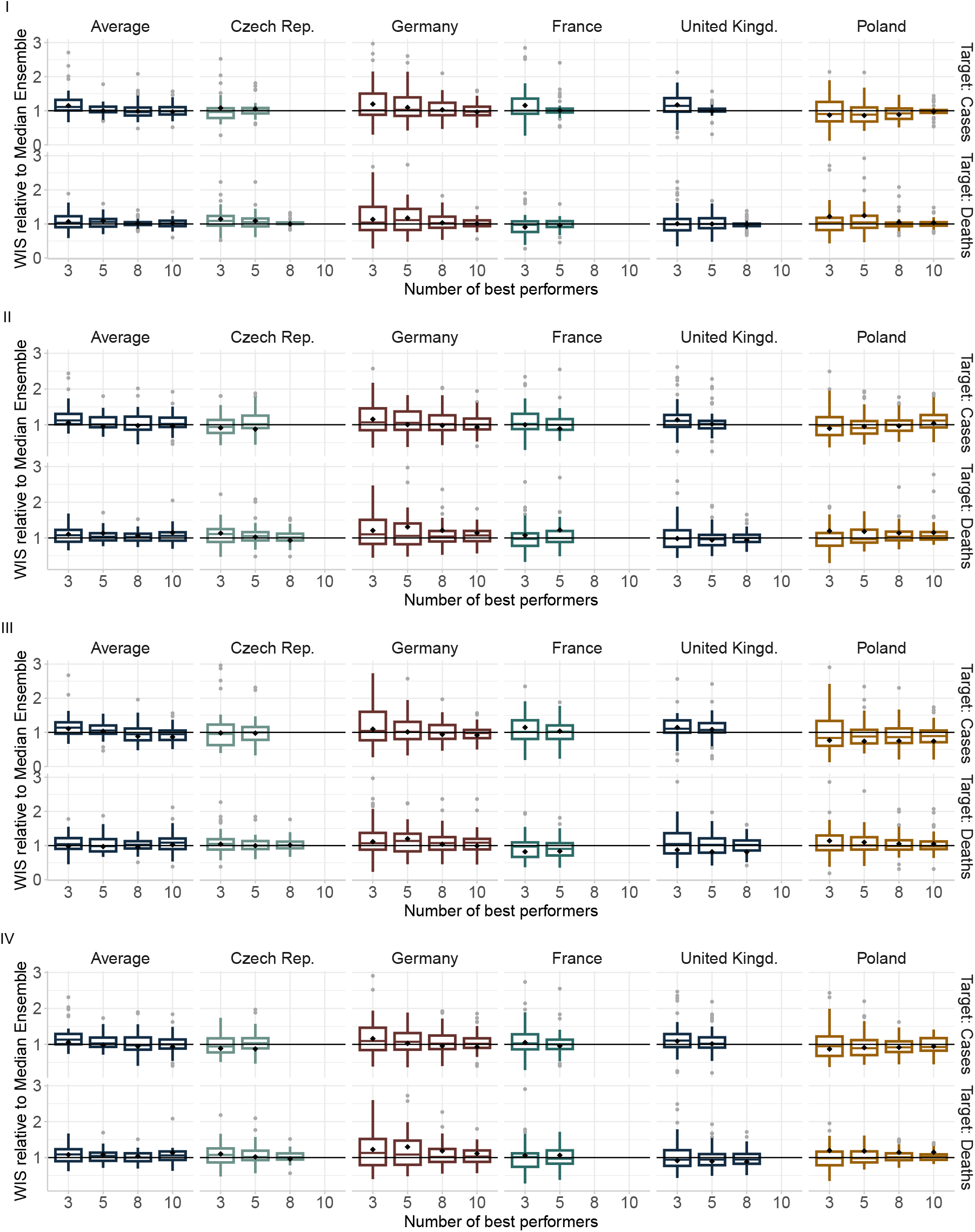
WIS spread of the selection ensemble methods relative to the Hub ensemble, by location, target series, and number of best recent performers (k) included. The four panels show the different aggregation methods. (I): equally weighted median. (II): equally weighted mean. (III): inverse score weighted median. (IV): inverse score weighted mean. For all methods, selection of component forecasts and (where applicable) estimation of weights for a given forecast origin was based on performance in the past four weeks. Outliers with relative scores over 3 were excluded from the plots for legibility - to compensate for this, the diamond-shaped black points show the mean relative score value. Values below one mean that the respective method outperformed the benchmark for the given target. Missing boxplots for a given k indicate that the corresponding location and target type combination lacked a sufficient model base to support the analysis.

## References

Amaral AVR, Wolffram D, Moraga P, and Bracher J (2025). “Post-processing and weighted combination of infectious disease nowcasts”. PLoS Computational Biology 21.3, e1012836.

Batchelor R and Dua P (1995). “Forecaster diversity and the benefits of combining forecasts”. Management Science 41.1, 68–75.

Bates JM and Granger CWJ (1969). “The Combination of Forecasts”. OR 20.4, 451–468.

Bosse N, Gruson H, Cori A, van Leeuwen E, Funk S, and Abbott S (2024). “Evaluating Forecasts with scoringutils in R”. Preprint, https://arXiv.org/abs/2205.07090.

Bracher J, Ray EL, Gneiting T, and Reich NG (2021a). “Evaluating epidemic forecasts in an interval format”. PLOS Computational Biology 17.2, e1008618.

Bracher J, Wolffram D, Deuschel J, Görgen K, Ketterer JL, Ullrich A, et al. (2021b). “A pre-registered short-term forecasting study of COVID-19 in Germany and Poland during the second wave”. Nature Communications 12.1, 5173.

Claeskens G, Magnus JR, Vasnev AL, and Wang W (2016). “The forecast combination puzzle: A simple theoretical explanation”. International Journal of Forecasting 32.3, 754–762.

Cramer EY, Ray EL, Lopez VK, Bracher J, Brennen A, Castro Rivadeneira AJ, et al. (2022). “Evaluation of individual and ensemble probabilistic forecasts of COVID-19 mortality in the United States”. Proceedings of the National Academy of Sciences 119.15, e2113561119.

Fox SJ, Kim M, Meyers L, Reich NG, and Ray EL (2024). “Optimizing Disease Outbreak Forecast Ensembles”. Emerging Infectious Diseases 30.9, 1967–1969.

Grieve R, Yang Y, Abbott S, Babu GR, Bhattacharyya M, Dean N, et al. (2023). “The importance of investing in data, models, experiments, team science, and public trust to help policymakers prepare for the next pandemic”. PLOS Global Public Health 3.11, e0002601.

Johansson MA, Apfeldorf KM, Dobson S, Devita J, Buczak AL, Baugher B, et al. (2019). “An open challenge to advance probabilistic forecasting for dengue epidemics”. Proceedings of the National Academy of Sciences 116.48, 24268–24274.

Knutti R, Furrer R, Tebaldi C, Cermak J, and Meehl GA (2010). “Challenges in Combining Projections from Multiple Climate Models”. Journal of Climate 23.10, 2739–2758.

Lamberson PJ and Page SE (2012). “Optimal Forecasting Groups”. Management Science 58.4, 805–810.

McGowan CJ, Biggerstaff M, Johansson M, Apfeldorf KM, Ben-Nun M, Brooks L, et al. (2019). “Collaborative efforts to forecast seasonal influenza in the United States, 2015–2016”. Scientific reports 9.1, 683.

Ray EL, Brooks LC, Bien J, Biggerstaff M, Bosse NI, Bracher J, et al. (2023). “Comparing trained and untrained probabilistic ensemble forecasts of COVID-19 cases and deaths in the United States”. International Journal of Forecasting 39.3, 1366–1383.

Reich NG, Lessler J, Funk S, Viboud C, Vespignani A, Tibshirani RJ, et al. (2022). “Collaborative Hubs: Making the Most of Predictive Epidemic Modeling”. American Journal of Public Health 112.6, 839–842.

Resin J, Wolffram D, Bracher J, and Dimitriadis T (2024). “Shift-Dispersion Decompositions of Wasserstein and Cramér Distances”. Preprint, https://arxiv.org/abs/2408.09770.

Sherratt K, Grah R, Prasse B, Becker F, McLean J, Abbott S, et al. (2025). “The influence of model structure and geographic specificity on predictive accuracy among European COVID-19 forecasts”. Preprint, https://www.medrxiv.org/content/10.1101/2025.04.10.25325611v1.

Sherratt K, Gruson H, Grah R, Johnson H, Niehus R, Prasse B, et al. (2023). “Predictive performance of multi-model ensemble forecasts of COVID-19 across European nations”. eLife 12, e81916.

Sherratt K, Gruson H, Johnson H, Niehus R, Prasse B, Sandman F, et al. (2022). European Covid-19 Forecast Hub. Dataset, Zenodo: 10.5281/zenodo.7669867.

Viboud C, Sun K, Gaffey R, Ajelli M, Fumanelli L, Merler S, et al. (2018). “The RAPIDD ebola forecasting challenge: Synthesis and lessons learnt”. Epidemics. The RAPIDD Ebola Forecasting Challenge 22, 13–21.

